# The Case for Interpretable Geometry: Statistical Shape Models vs. Curvature-Based Descriptors in Aortic Disease Classification

**DOI:** 10.64898/2026.07.29.26359299

**Authors:** Duc Nguyen, Joseph A. Pugar, Luka Pocivavsek

## Abstract

**Purpose:** Quantifying aortic morphology is central to surgical planning for thoracic endovascular aortic repair (TEVAR), yet no consensus exists on how best to represent three-dimensional aortic shape for outcome prediction. Two broad strategies have emerged: statistical shape analysis (SSA), which relies on statistical methods and dimensionality reduction to capture the most significant shape modes, and geometrically-informed approaches that extract descriptors grounded in differential geometry. Here, we directly compare these paradigms on a cohort of 290 CTA scans classified by surgical outcome (non-pathological, successful TEVAR, failed TEVAR).

**Methods:** For the geometrically-informed approach, we use a two-dimensional feature space using normalized fluctuation in integrated Gaussian curvature 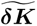 and mean aortic radius ***R***. For SSA, we construct a point-cloud shape model with dimensionality reduction using Principal Component Analysis (PCA) and evaluate classification performance as a function of the number of retained principal components.

**Results:** SSA’s leading principal components encode variations in global aortic size and are statistically redundant with (***R***, 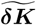), yet they lack a one-to-one correspondence with interpretable anatomical quantities. Testing on an unseen, independent dataset reveals that the geometrically-informed approach provided better generalizability than SSA. Using Gaussian process classification with 10-fold cross-validation, we find that the geometrically-informed approach achieves a higher weighted ***F*_1_** score than SSA achieves with up to 20 principal components. While SSA’s full-dataset accuracy rises above 90% with increasing dimensionality, this gain is driven by overfitting rather than genuine discriminative power.

**Conclusion:** These results demonstrate that geometrically-informed descriptors offer a more interpretable, robust, and clinically translatable framework for aortic disease classification than data-driven statistical shape representations.

## 1 Introduction

Vascular surgeons have long recognized that anatomical morphology guides both the feasibility and success of endovascular intervention. In thoracic endovascular aortic repair (TEVAR), patient selection and device planning depend heavily on geometric assessment of the aorta, yet the features most commonly measured, such as maximal diameter, landing zone length, and arch angulation, represent only a small subset of the information encoded in three-dimensional aortic shape. Geometric features have proven valuable across cardiovascular medicine: aortic diameter thresholds inform prophylactic repair decisions [1], centerline tortuosity correlates with access complications and endoleak risk [2], and arch morphology classification schemes attempt to stratify procedural difficulty [3]. However, quantifying the shape of the aorta remains a challenge. Generally, two broad strategies have emerged for the featurization of aortic shape prior to downstream modeling: data-driven statistical representations that parametrize shape through dimensionality reduction techniques, and the calculation of geometric quantities grounded in differential properties of the aortic surface. Each offers distinct trade-offs between model interpretability and the ability to characterize complex morphology.

Curvature provides a natural framework for describing the aortic surface, motivating a geometric approach based on curvature-derived metrics. Among these metrics, Gaussian curvature *K*, an inherent measure of local surface geometry, emerges as a highly relevant descriptor. Additionally, prior studies have demonstrated that local curvature measurements can be aggregated into global geometric variables to characterize aortic morphology and support disease classification [4]. Building on these two ideas, we want to aggregate local, varying Gaussian curvatures into a global variable that characterizes overall aortic shape. By the Gauss–Bonnet theorem [5], the total curvature ∑ *K* = 0 over a closed aortic surface and remains invariant under surface deformation. Given this invariance, the spatial variability of curvature, quantified by the area-weighted fluctuation *δK*, serves as an effective descriptor. *δK* provides a size-invariant measure of geometric complexity of the entire aortic surface, where larger values indicate greater heterogeneity in surface curvature. Previous work by our lab has shown that this fluctuation in total curvature, *δK*, is a reliable shape descriptor for predicting whether a patient achieved successful or failed outcomes after TEVAR [6]. Subsequently, we showed that with a motivated choice of meshing and partitioning parameters, *δK*, along with mean root Casorati curvature as the size descriptor, can also be able to predict postoperative TEVAR outcome with an accuracy of 83% based only on preoperative information [7].

In addition to relying on well-defined geometrical quantities, there has been substantial literature that applied statistical shape analysis (SSA) to explore new and data-driven aortic shape representations [2, 8–12]. Within these approaches, a mean shape is often constructed from an aligned set of shapes, and each shape in this set is represented by its difference to the mean shape according to a certain metric, either through point-to-point correspondence (point distribution approach) or through how the template shape can be deformed to the shape under investigation (shape deformation approach). The point distribution approach quantifies a shape by points called landmarks on the shape’s boundary, which can be correlated between shapes through known anatomical understanding or parametric descriptors. Both methods of creating landmarks have been applied widely in clinical settings [13, 14]. The shape deformation approach, on the other hand, quantifies a shape through the diffeomorphic transformation from the template shape to the shape under investigation. The shape deformation approach has been applied to assess arch morphology [15] and bicuspid aortic valve and aortic coarctation [16]. In both point distribution and shape deformation approaches, dimensionality reduction is needed to extract the most useful shape features for analysis. This is often done by Principal Component Analysis (PCA) [17], which finds the vector directions that explain the majority of variance in a high-dimensional dataset. As noted by Zhang et al. [18], other non-linear methods of dimensionality reduction are sometimes used as the shape matrix can be highly non-linear.

Our geometric approach using 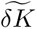, while interpretable and reliable, might not capture every aspect of the shape space. In addition, calculations of 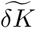 rely upon differential geometry, a mathematical framework that may be less immediately accessible to researchers outside the mathematics community, including surgeons and biophysicists. On the other hand, while SSA might be able to explore a more data-optimized shape representation, this approach has been criticized by lack of consistency and interpretability [19, 20]. This paper aims to provide a direct comparison between these two approaches on classification effectiveness. To do so, we will develop and apply SSA on our dataset using a point cloud based approach and compare the results of both geometric and SSA approaches on classifying whether an aorta belongs to a normal patient, a successful TEVAR intervention or a failed TEVAR intervention. Moreover, we will investigate the interpretability and generalizability of the SSA approach to conclude about the method’s applicability in a wider setting.

## 2 Methods

### 2.1 Clinical data collection, segmentation, and meshing

290 scans from 175 patients are selected for SSA and geometrically-informed approach. Three possible outcomes are assigned to the trimmed dataset: normal aortas (163 scans from 135 patients), successful TEVAR (50 scans from 18 patients), and failed TEVAR (77 scans from 22 patients). The criteria for labeling patients and their corresponding scans follow our previous analyses [6, 7]. All data collection and analysis are performed following the guidelines established by the Declaration of Helsinki and under institutional review board approval (IRB20-0653, IRB21-0299). Data are received as deidentified DICOM files from CTA instrumentation spanning a variety of scanners and resolutions (in-plane spacings from 0.3–0.8 mm and slice thicknesses from 0.3– 3.0 mm). Segmentation of the aortic lumen and centerline extraction is performed semi-automatically using PraevAorta software developed by Nurea [9, 21]. Segmented surfaces are exported as triangulated meshes with an extrinsic element size of 2.5mm^2^ to balance computational efficiency with geometric fidelity.

### 2.2 Geometrically-informed aorta morphology extraction

The geometrically-informed approach extracts shape and size descriptors grounded in differential geometry, following the framework developed by our lab previously [6] and refined through scale-space optimization [7]. For each aortic surface mesh, discrete Gaussian curvature *k_g_* is estimated per vertex using a finite-difference algorithm [22]. The mesh is then partitioned into *k* patches, and the integrated Gaussian curvature within each patch is computed as *K_j_* = *A_j_*⟨*k_g_*⟩, where *A_j_* is the patch area and ⟨*k_g_*⟩ is the area-weighted mean Gaussian curvature within the patch. The shape descriptor *δK* captures the fluctuation in these per-patch curvature integrals:

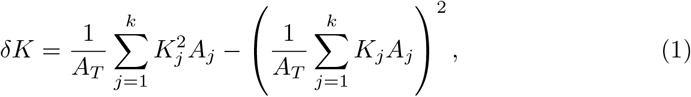

where *A_T_* is the total aortic area. The size descriptor in the size-shape feature pair is either radius (in the earliest work inverse radius) or aortic surface area. The values of *δK* depend on preprocessing choices—specifically, the degree of surface smoothing, mesh element density, and partition size. Following Pugar et al. [7], we adapt scale-space parameters from the “stable zone”, yielding patch areas on the order of 1−2cm^2^. *δK* is normalized to the non-pathological cohort mean, yielding 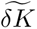, which is used alongside the mean aortic radius *R* to define the latent anatomic feature space used for classification tasks.

### 2.3 Landmark generation and principal component analysis

For the SSA approach, we follow the implementation by Wiputra et al. [23], which uses the normalized length of the aorta *s* and polar angle *θ* as the aorta’s parametric coordinate system on which generation of landmark points relied. Our implementation of SSA can be summarized as:

1. The aortas and centerlines are exported using a common coordinate system, which are then centered, normalized using the centerline length, and aligned using the Procrustes method.
2. We sample *N_s_* points equally spaced along the arclength centerline.
3. We calculate the tangent vectors at each chosen centerline point using a sliding window average. We use a window size of 5 points to smooth the tangent calculation.
4. For each centerline point and tangent, we define a plane with the tangent normal. We sample *N_θ_* points, equally spaced by angle, from the curve generated by the intersection of the aorta surface with the plane. The detailed landmark sampling algorithm (Algorithm 1) can be found in Appendix A. Thus, we have *N_s_* × *N_θ_* landmarks that will parameterize the aorta. For the rest of the analysis, we take *N_s_* = *N_θ_* for equal sampling in both parameters. We then extract from those points the radius *r*, calculated by the distance from the point to the corresponding centerline point.
5. We build a matrix of the form 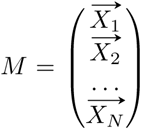, with 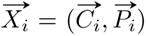. 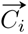 contains the coordinates of each centerline point (*x, y, z*) extracted for scan *i*. 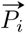 contains all of the radii of the sampled points along the aorta. Both 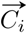 and 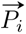 has been normalized across aorta populations.
6. We perform PCA on the matrix *M* using scikit-learn [24] PCA functionality. For equal comparison to the geometrically-informed approach, each feature is normalized to the non-pathological cohort mean.

We implement logistic regression (LR) and Gaussian process classification (GPC) to determine whether a scan is normal, failed TEVAR, or successful TEVAR by learning the boundaries within the feature space. LR learns linear decision boundaries, and is used in our analysis to examine how classification performance varies with number of input features and principal components retained in the SSA approach. Head-to-head comparison between geometric and SSA approaches in two-dimensional space utilizes GPC with a radial basis function (RBF), as its ability to model non-linear decision boundaries provides greater flexibility and therefore enables comparison under the best achievable classification performance for both approaches. For the geometric approach, the regression inputs are *R* and 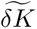. For the SSA approach, the inputs are normalized scores in the principal component space. Classification accuracy is determined by the weighted *F*_1_ score. A visualization of the SSA pipeline, in comparison to the geometrically-informed pipeline, is shown in Figure 1.

**Fig. 1.**
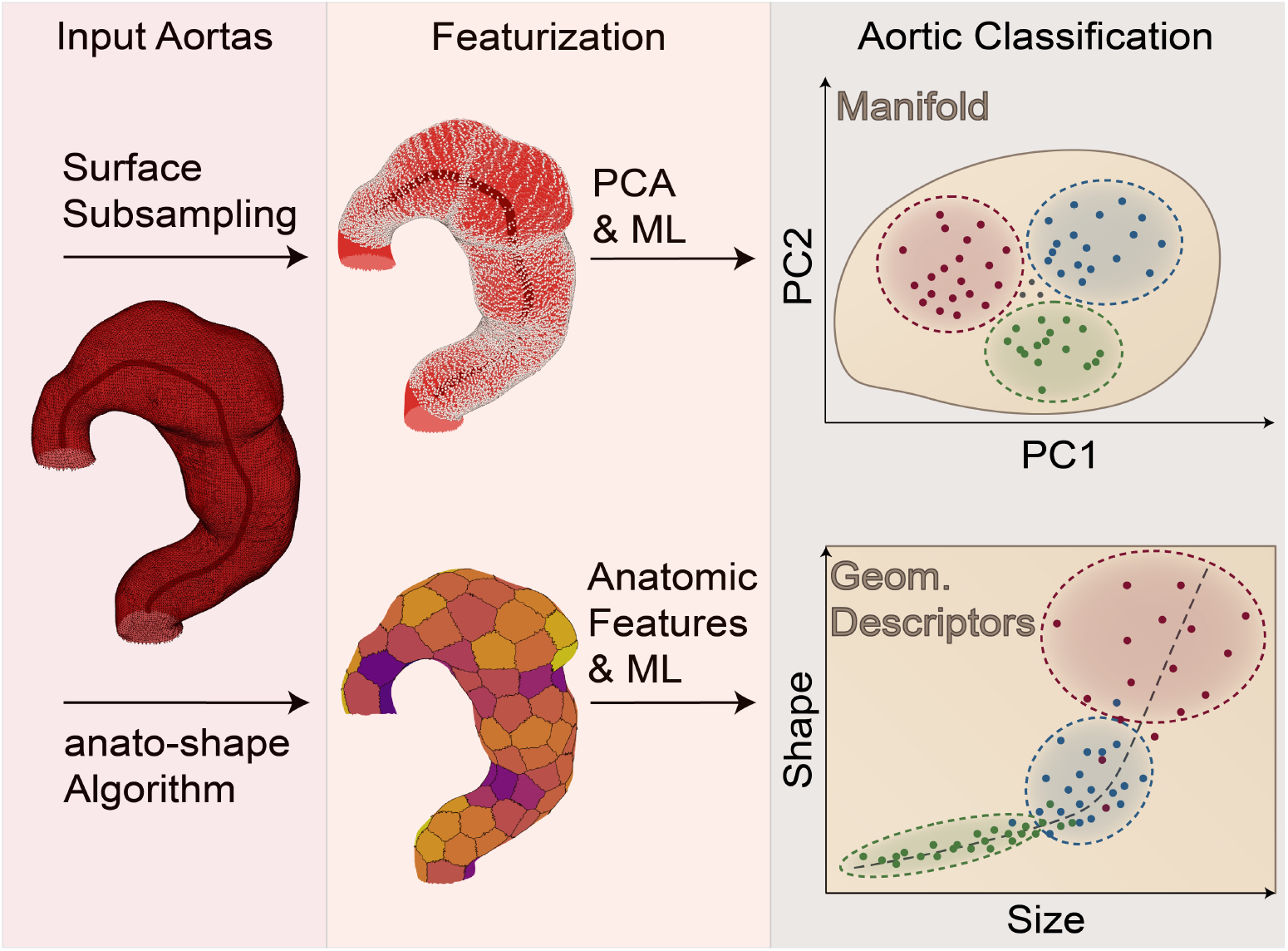
Overview of the two featurization strategies compared in this study. Both pipelines begin with patient-specific aortic surface meshes reconstructed from CTA scans. In the statistical shape analysis (SSA) approach, each mesh is uniformly subsampled along centerline cross-sections, and the resulting high-dimensional radius vectors are reduced via principal component analysis (PCA). The leading principal component (PC) scores are then passed to a classifier operating in the manifold. In the geometric approach, anatomically grounded features of size and shape are extracted from the meshes (i.e., *R* and 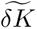) that serve as direct inputs to a classifier in a physically interpretable feature space. Both pathways terminate in a three-class aortic classification task (Normal, TEVAR Success, TEVAR Failure).

## 3 Results

### 3.1 SSA results

Ensuring alignment across aortas is crucial in SSA to confirm that the shape model captures local shape differences rather than rotational and translational transformations. To quantify alignment, we use the Surface DICE metric [25]. In brief, Surface DICE evaluates the degree of agreement between two surfaces (here, aortic geometries) by assessing how closely the surfaces overlap within a selectable distance tolerance, *τ*. Specifically, the metric computes the proportion of surface points from each geometry that lie within a distance *τ* of the other surface. A score approaching 1 indicates a high degree of overlap and, consequently, strong alignment between the two surfaces. Figure 2 shows the alignment quality of aortas in the cohort after preprocessing and before landmark generation (after step 1 of Section 2.3). In Figure 2**(A)**, as the tolerance threshold increases, the Surface DICE score also increases and approaches 1. This indicates that the aortas are properly aligned: lower scores at smaller tolerances reflect sensitivity to local shape differences, while scores approaching 1 at larger tolerances indicate that the major anatomical features (such as the aortic arch and ascending/descending segments) are well aligned. Alignment of aortas can also be seen visually in **Figure 2(B)** and **(C)**.

**Fig. 2.**
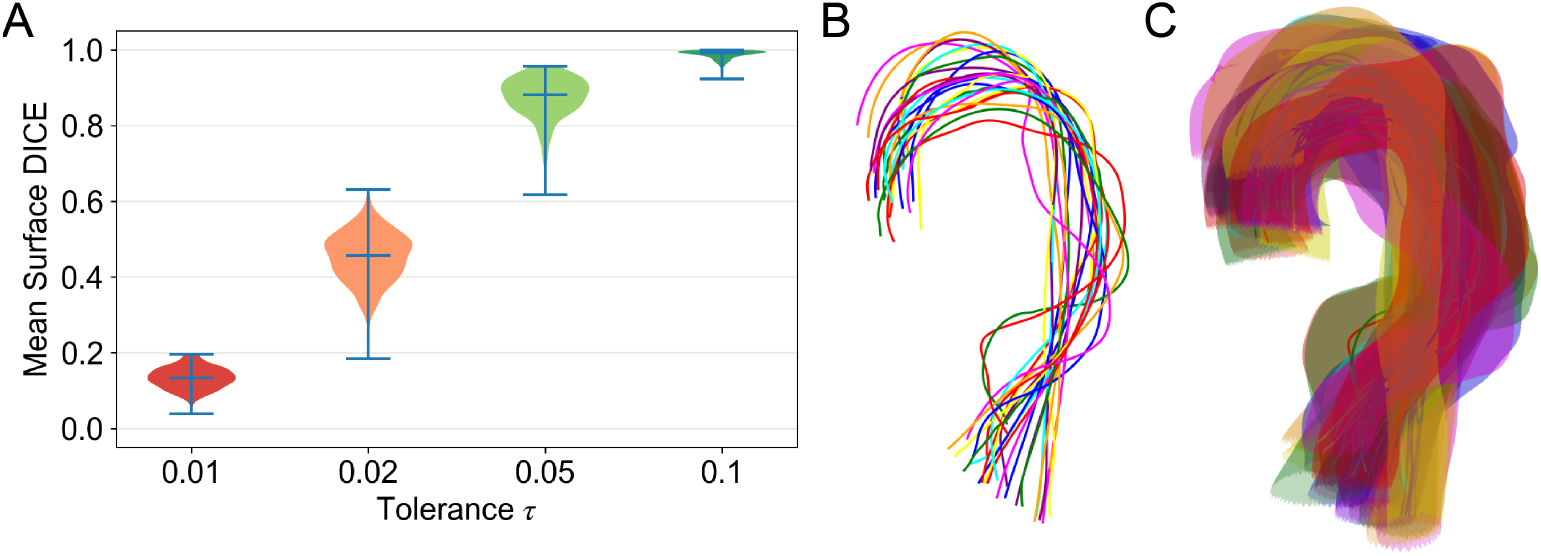
Mesh alignment quality and cohort registration. **(A)** Violin plots of per-subject mean Surface DICE scores computed over 20 random pairwise comparisons (1 000 subsampled points per mesh) at four tolerance thresholds *τ* ∈ {0.01, 0.02, 0.05, 0.1}. Horizontal bars indicate the median. At *τ* = 0.1, the median Surface DICE approaches 1.0, confirming that the Procrustes-based registration brings meshes into close spatial agreement. **(B)** Overlay of 25 random aligned centerlines after centering, arc-length scaling, and Procrustes rotation, illustrating the residual anatomical variability in aortic arch curvature across subjects. **(C)** Superposition of the same set of aligned surface meshes, demonstrating the range of aortic morphology captured by the cohort.

We also investigate the impact of sampling density on classification performance and find that varying the number of sampled points has an effect on classification results. As shown in Figure 3**(B)**, the highest classification accuracy with multinomial LR is 87.1±0.4%, achieved when *N_s_* = *N_θ_* = 10 (130 input features). In our case, SSA doesn’t require a large number of points; the weighted *F*_1_ score decreases as the number of points sampled increase. However, Figure 3**(A)** demonstrated that increasing the number of sampled points can improve the efficiency of the representation, as a greater proportion of the total variance is captured by the first few principal components, indicating that these components encode more of the underlying shape information.

**Fig. 3.**
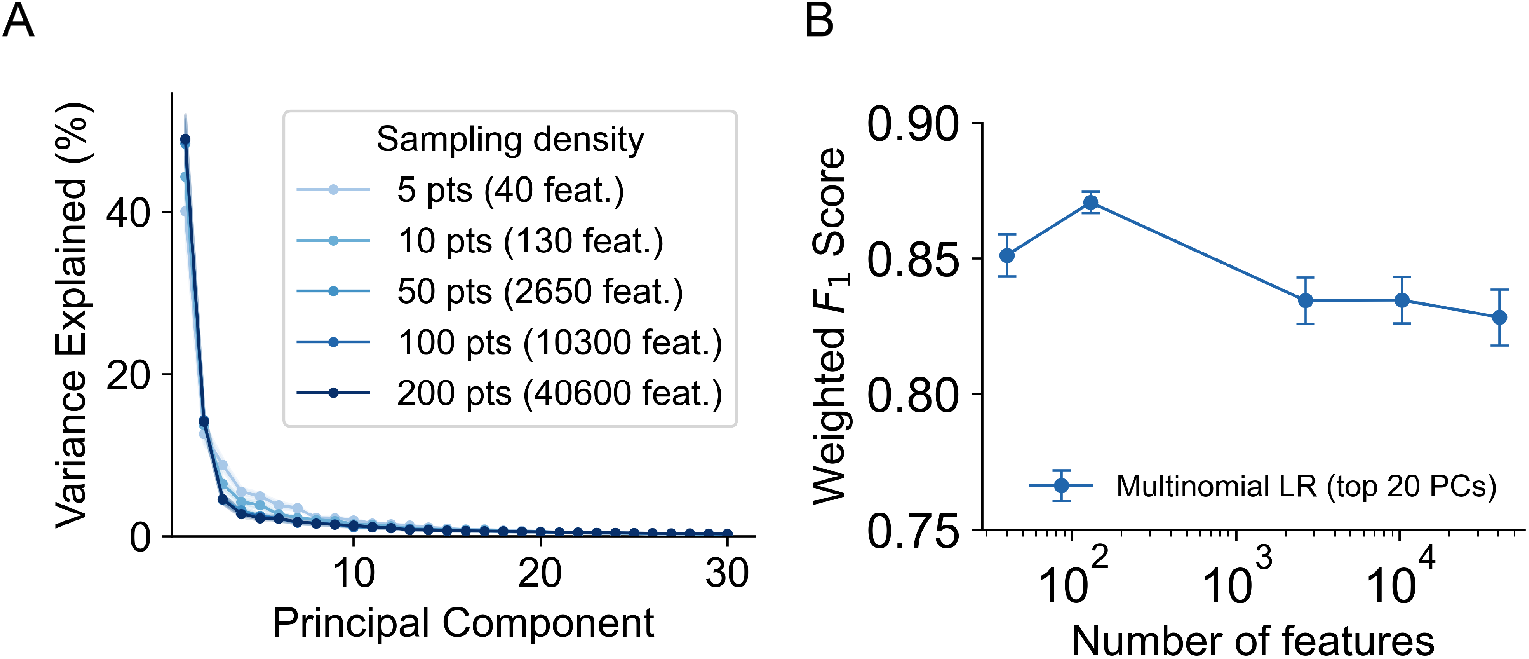
Effect of surface sampling density on PCA and classification. **(A)** Variance explained by the first 30 principal components at five sampling densities: *N_s_* = *N_θ_* = 5, 10, 50, 100, and 200 points (corresponding to 40; 130; 2650; 10300; and 40600 total features, respectively). Shaded bands denote *±*1 standard deviation across 10 bootstrap resamples. Lower sampling densities spread variance across more components, with diminishing marginal gains beyond *∼*10 PCs at all resolutions. **(B)** Weighted *F*_1_ score from multinomial logistic regression (LR) using the top 20 PCs, plotted against the number of input features. Classification performance peaks near 130 features (10 pts) and declines at higher resolutions.

A closer look into the distribution of the aorta dataset in the principal component space can be found in Figure A4 in Appendix A section. In the space formed by the first two principal components (PC_1_ vs PC_2_), there is a clear separation between normal versus successful and failed TEVAR. The decision boundary between successful and failed TEVAR is less clear. However, there is no noticeable separation between the different outcomes in the feature space formed by PC_3_ and PC_4_. These principal components are not useful in our goal of outcome classification.

### 3.2 Comparison to geometrically-informed approach

Figure 4 shows the performance comparison of both approaches using GPC applying on the entire cohort. For the same number of classification dimensions (2 principal components versus (*R*, 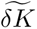)), SSA approach produces a lower *F*_1_ score than the geometric approach (87.8% compared to 90.5%). A paired bootstrap analysis on this cohort (Figure A1 A) indicates that, on average, the SSA-based approach yields lower performance across resampled datasets compared to the geometrical approach. However, this observed difference in weighted *F*_1_ score does not reach statistical significance at the *p <* 0.05 level, as the 95% confidence interval includes zero (95% CI: [−10.24%, 3.98%]). When trained on a linear classifier (Figure A2 A), the *F*_1_ scores of the SSA approach is only slightly lower than that of the geometric approach (85.2% versus 86.5%). Visually, the geometric approach clearly separates normal, successful and failed aortas into distinct bands in the (*R*, 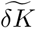) space. On the other hand, the SSA approach results in overlapping class regions, with several aortas clustering near the intersecting decision boundary of three outcomes, leading to classification ambiguity.

**Fig. 4.**
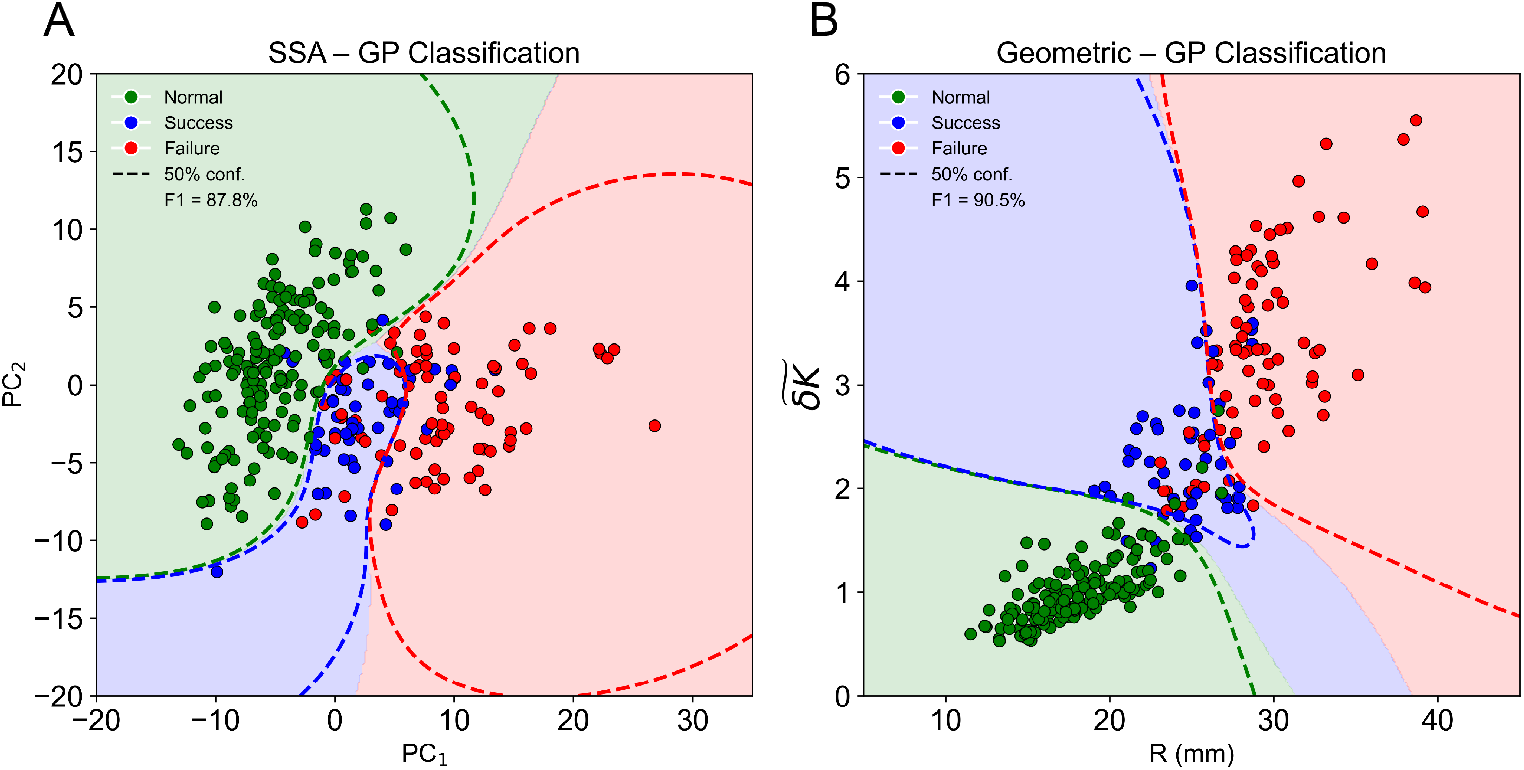
Gaussian process classification decision boundaries. **(A)** SSA feature space: Gaussian process classifier (GPC) with a radial basis function (RBF) kernel trained on the first two principal components (PC_1_ vs. PC_2_). Shaded regions indicate predicted class territories; dashed contours mark the 50% posterior probability boundary for each class (weighted *F*_1_ = 87.8%). **(B)** Geometric descriptor feature space: GPC with an RBF kernel trained on (*R* vs. 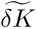). The geometric model achieves a higher *F*_1_ of 90.5% with decision boundaries that separate the three outcome classes along axes with direct anatomical meaning: larger, more irregular aortas cluster toward the Failure region. Green: Normal; blue: TEVAR Success; red: TEVAR Failure.

One might argue that the power of principal component analysis lies in its ability to incorporate more than 2 dimensions to assist with classification. Indeed, as the number of retained principal components increases, classification accuracy can reach over 90%, the red line in Figure 5**(B)**, which approaches the optimal geometric results. This raises the question of whether the performance gain reflects intrinsic discriminative power or reliance on high-dimensional aggregation, which we examine by comparing *k*-fold accuracies for SSA approach across multiple principal components to the geometrically informed approach. *k*-fold is a more robust and reliable way to assess the algorithm’s performance. By splitting the data into folds and using every fold as both training data and testing evaluation, we can prevent model overfitting and ensure generalization of the model to unseen data. In detail, we train the dataset on *k*−1 folds and calculate the testing accuracy on the leftover fold using weighted *F*_1_ score. We repeat this process *k* times, each time leaving out a different fold. *k*-fold cross-validation is done using scikitlearn [24], and we set *k* = 10. As shown in Figure 5**(A)**, the weighted *F*_1_ score for the SSA approach peaks at *N_p_* = 20 (*F*_1_ score = 87%), higher than the original geometric approach (*F*_1_ score = 85%). The geometrically-informed approach with scale space optimization (blue dashed line) outperforms both the original geometric approach or SSA.

**Fig. 5.**
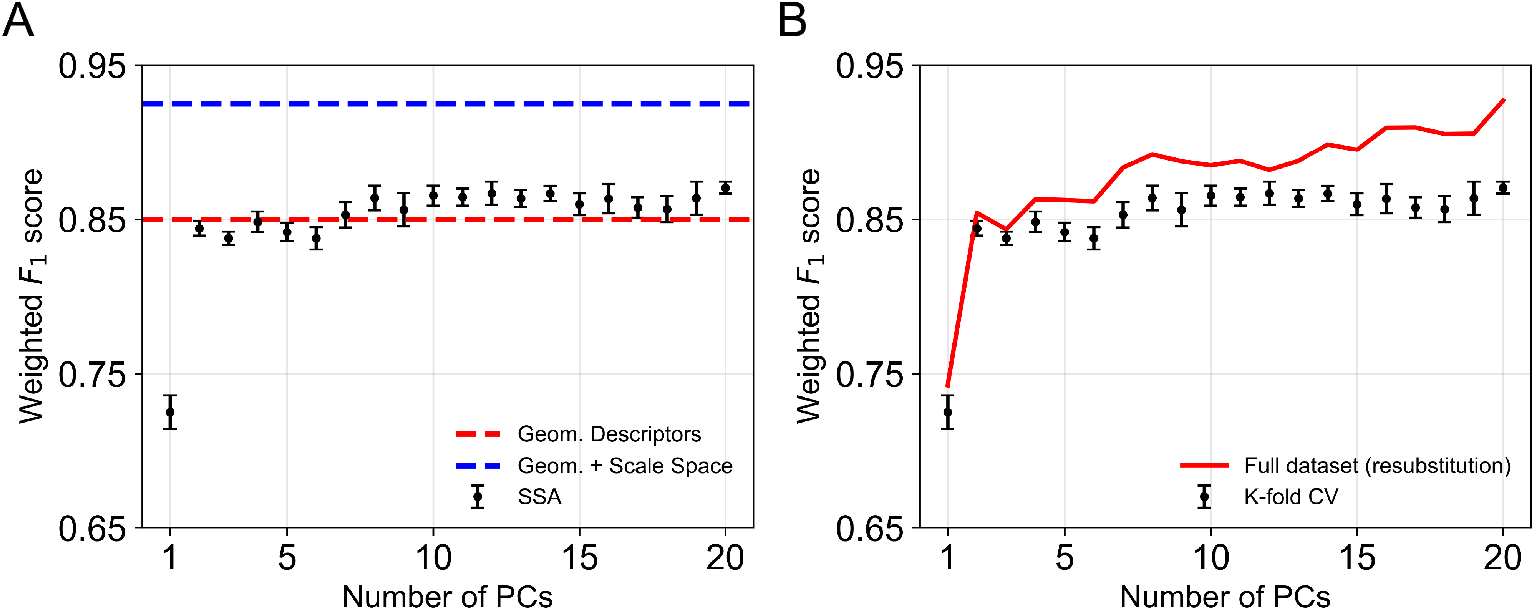
Classification performance of SSA versus geometric descriptors. **(A)** Weighted *F*_1_ score of multinomial logistic regression (LR) as a function of the number of retained principal components (*N_p_* = 1–20). Each black point represents the mean over 10 repetitions of 10-fold cross-validation; error bars denote *±*1 standard deviation. The red dashed line marks the *F*_1_ achieved by the original geometric descriptor model (*R*, 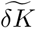), and the blue dashed line marks the *F*_1_ of the geometric model augmented with scale-space optimization [7]. SSA performance plateaus near *N_p_ ≈* 10 and slightly exceeds the geometric descriptor baseline. **(B)** Overfitting diagnostic. The red curve shows the resubstitution (full-dataset) *F*_1_, which rises steadily with additional PCs, while the cross-validated *F*_1_ (black points) remains flat beyond *N_p_ ≈* 3. The widening gap indicates that additional PCs capture noise rather than generalizable signal.

A comparison between the accuracy when training and testing on the entire dataset versus on *k*-folds in Figure 5**(B)** reveals why this is the case. For the first two principal components, the accuracy obtained using the full dataset is consistent with the *k*-fold cross-validation results. As additional principal components are retained, however, a divergence emerges: the full-dataset accuracy increases to over 90%, whereas the *k*-fold accuracy remains at 85–87%. This pattern is characteristic of overfitting, where improvements in training performance don’t match with similar improvements in testing accuracy.

## 4 Discussion

### 4.1 Interpretability of SSA results

As explored in the previous section, the first few principal components hold the most information useful for outcome classification. This section aims to understand the correlation between the principal components and known geometric descriptors—mean aortic radius and integrated Gaussian curvature.

Figure A5 (A) shows the relationship between principal components and mean aortic radius. The first principle component shows a strong positive correlation with mean aortic radius (*r* = 0.80), while the second principal component shows a weak negative correlation (*r* = −0.44). We also test statistical shape analysis’s prediction on ideal shapes (bent cylinders of increasing radius with no curvature, 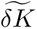 = 0) and find that the first two principal components are proportional to the radii of the ideal shapes, confirming this hypothesis. As Wiputra et al. [23] concluded, the first few principal components revealed information about size. However, the observed correlation strengths (*r* = 0.80 and −0.44) indicate that while PC_1_ functions as a robust and interpretable proxy for overall size, PC_2_ reflects a more entangled mixture of size and shape variations, lacking a clear or physically meaningful interpretation. It is also difficult to disentangle shape and size indicators, as it is known from our analysis of *R* and 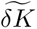 that these two size and shape variables are positively correlated. Figure A6 (A) demonstrates this point, as PC_1_ is also correlated to 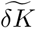 (*r* = 0.83). We also find no correlation between PC_3_ and PC_4_ to aortic radius and integrated Gaussian curvature, suggesting that further principal components are dominated by irrelevant shape modes. We note that some prior work has reported interpretable SSA modes in other aortic pathologies; for example, Cosentino et al. [8] found that principal shape modes of ascending thoracic aortic aneurysms correspond to identifiable features such as vessel tortuosity and local bulging. The lack of comparable interpretability in our TEVAR cohort suggests that the mapping from PC modes to anatomically meaningful features is not guaranteed and depends on the specific disease context and shape representation.

### 4.2 Generalizability of SSA results

To test if SSA’s classification can generalize to unseen data, we run the SSA pipeline on a new dataset, which contains 152 scans from patients belonging to the GORE TAG 08-01 dataset [26] who underwent TEVAR using a stent graft for type B aortic dissection. This dataset is processed using the same meshing algorithm as the original dataset, as described in Section 2.1. We then use the principal component model and the Gaussian process classifier trained earlier to produce predictions (normal or pathological) on this GORE TAG 08-01 dataset.

We expect an approach which has captured the shape variations necessary for disease classification and can be generalized to new datasets to be able to classify the scans in this unseen dataset as pathological, corresponding to the fact that the patients underwent TEVAR for aortic injuries. However, as shown in Figure 6, SSA classifies half of scans as normal, significantly higher than the geometric method (23.2%). This limited generalizability to a similar dataset reinforces our concern about the lack of interpretability of SSA. It remains unclear whether the modes of greatest shape variation found by SSA (PC_1_ and PC_2_) truly encapsulate clinically meaningful information and demonstrate its limitations.

**Fig. 6.**
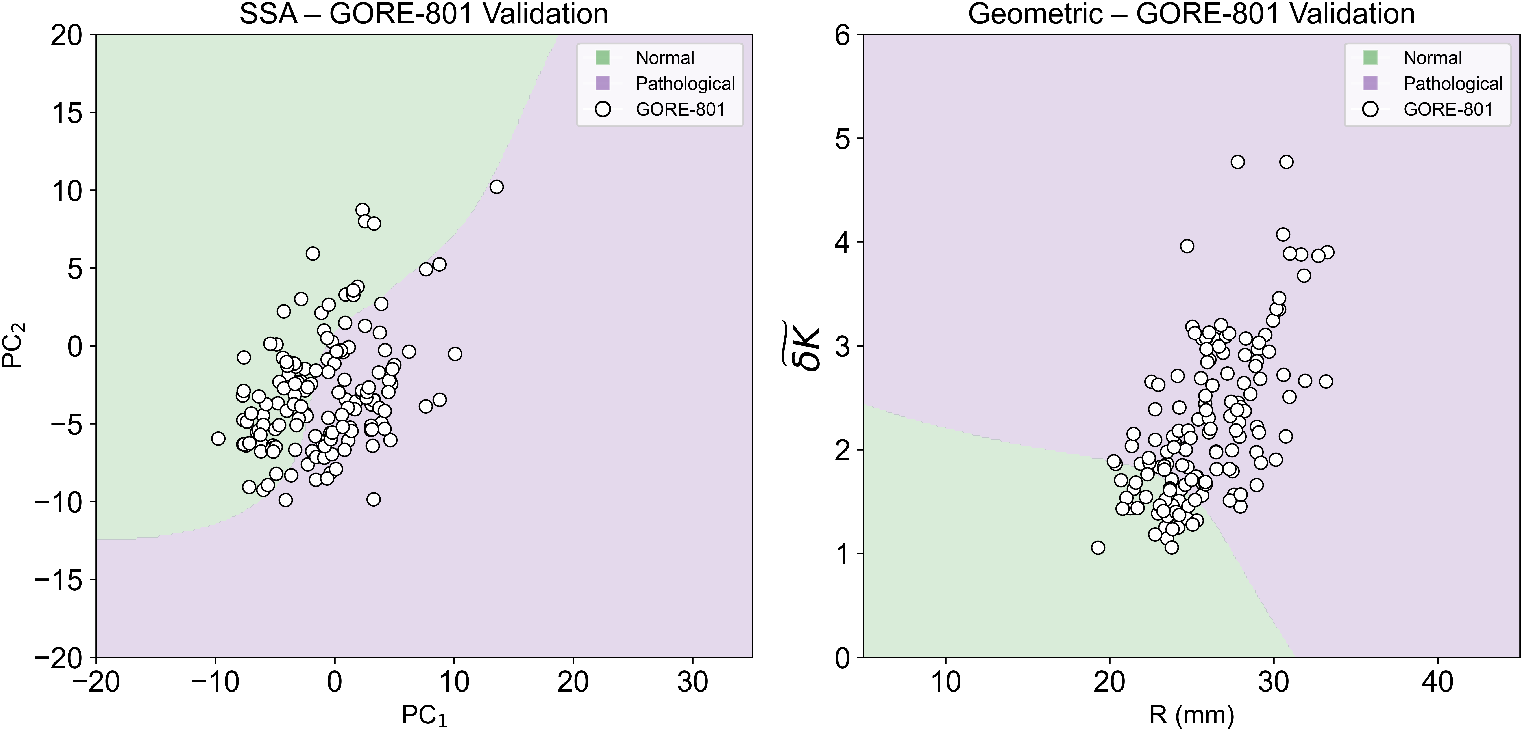
Generalizability assessment on an independent dataset. Both classifiers (GPC with RBF kernel, trained on the original 290-scan cohort) are applied to 152 unseen CTA scans from the GORE TAG 08-01 trial [26], comprising patients who underwent TEVAR with a conformable stent graft for acute, complicated type B aortic dissection. Because every patient in this cohort required endovascular intervention, at the cohort level, scans are expected to be classified as pathological, although a few individual scans which reflect post-repair modeling towards normal geometry may appear morphologically borderline. **(A)** In the SSA feature space (PC_1_ vs. PC_2_), 50.0% of GORE TAG scans fall within the normal region, indicating that the principal component decision boundary does not generalize to an independent population. **(B)** In the geometric descriptor feature space (*R* vs. 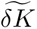), only 23.2% of scans are classified as normal. Decision boundaries are similar to Figure 4, with TEVAR success and TEVAR failure grouped together as pathological (purple region). The substantially lower normal-misclassification rate of the geometric approach supports its superior generalizability and reinforces the concern that SSA’s leading principal components do not reliably encode clinically meaningful shape variation across cohorts.

## 5 Conclusion

The result that PC_1_ and PC_2_ hold the most information that is important for classification meant that both these methods essentially relied on a 2-dimensional feature space in order to predict the outcome of each patient. In two-dimensional feature space predictions, the geometric method performs better than SSA. Furthermore, the lack of a clear association for PC_1_ and PC_2_ of SSA to concrete physical variables poses a major obstacle to interpret and contextualize our SSA results. On the other hand, the geometrically-informed approach is more interpretable and shows promise in extensibility to other clinical problems where shape is linked with disease state. The interpretability advantage of the geometric approach is not merely a matter of convenience, it reflects a fundamental difference in how each method relates to the underlying physics of aortic disease. The normalized fluctuation in integrated Gaussian curvature, 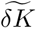, has a direct physical meaning: it quantifies the degree to which curvature is heterogeneously distributed across the aortic surface, a property that the Gauss–Bonnet theorem constrains and that correlates with the mechanical environment experienced by an endograft. A surgeon can reason about what a large 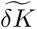 implies (e.g., regions of high local curvature that may compromise seal zones or promote endoleak) and can relate changes in 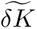 to specific anatomical features such as angulation at the arch or focal dilatation of the descending aorta. By contrast, a principal component score has no such transparent mapping. As our analysis in Section 4.1 demonstrated, PC_1_ and PC_2_ correlate with aortic size but do not isolate it cleanly, and they offer no direct insight into the spatial distribution of curvature or the biomechanical factors that drive device failure. For a tool to be adopted in surgical planning, clinicians must be able to understand not only what the model predicts but why, and geometric descriptors satisfy this requirement in a way that abstract statistical modes do not.

We finally conclude that our approach of using mathematically-proven shape and size descriptors is more interpretable and can achieve an accuracy comparable to, and with optimization, better accuracy for classifying outcome of patients. SSA is capable of capturing shape signals that can assist with classification tasks, as demonstrated by a strong classification performance (87.8%) when combined with a GPC. However, because of its lack of interpretability and generalizability, SSA is best applied to problems containing complex patterns after potential analytical or mathematical formulations have been thoroughly explored.

We note that SSA is a broad and diverse field, encompassing techniques such as shape deformation, non-parametric landmark sampling and non-linear approaches to dimensionality reduction. In this study, we evaluate one approach (i.e., point distribution model with linear PCA) and haven’t explored shape deformation models or non-linear embeddings of the shape space. Thus, our conclusions about model performance are restricted to point distribution models with linear PCA embedding. However, our message concerning interpretability and generalizability remains relevant: statistically derived shape modes do not necessarily correspond to anatomically meaningful features, nor do they inherently generalize to related clinical applications.

## Data Availability

The de-identified training cohort data (segmentation masks and derived shape descriptors) are publicly available at https://github.com/SurgBioMech/plos_data. The GORE TAG 08-01 validation dataset is not publicly available; access was provided to the University of Chicago through a data transfer agreement with W. L. Gore & Associates and is subject to the terms of that agreement.

## Supplementary information

Supplementary information accompanies this manuscript, including the landmark sampling algorithm, supplementary classification results, mesh alignment quality details, principal component space visualization, and correlation analyses between principal components and geometric descriptors (see Appendix A).

## Acknowledgements

The authors thank members of the SurgBioMech Lab at the University of Chicago for helpful discussions throughout the development of this work. The Center for Research Informatics is funded by the Biological Science Division, USA at the University of Chicago.

## Declarations

- **Ethical Approval:** This study was conducted in accordance with relevant laws, institutional guidelines, and the ethical principles outlined in the Declaration of Helsinki. Patient imaging data were obtained through the Human Imaging Research Office (HIRO) at the University of Chicago under Institutional Review Board (IRB) approvals IRB20-0653 (approved 7/15/2020) and IRB21-0299 (approved 3/24/2021).
- **Consent to Participate:** Informed consent was obtained from all participants, and the privacy rights of human subjects were observed throughout the study.
- **Consent to Publish:** All authors have approved the manuscript and consent to its publication.
- **Data Availability Statement:** The de-identified training cohort data (segmentation masks and derived shape descriptors) are publicly available at https://github.com/SurgBioMech/plos_data. The GORE TAG 08-01 validation dataset is not publicly available; access was provided to the University of Chicago through a data transfer agreement with W. L. Gore & Associates and is subject to the terms of that agreement.
- **Authors’ Individual Contributions:** Conceptualization: D.N., J.P., L.P. Data Curation: D.N., J.P. Formal Analysis: D.N. Funding Acquisition: L.P. Investigation: D.N. Methodology: D.N., J.P., L.P. Project Administration: L.P. Resources: L.P. Supervision: L.P. Validation: D.N. Writing – Original Draft: D.N., J.P. Writing – Review & Editing: D.N., J.P., L.P.
- **Funding:** This work was supported by the National Institutes of Health, USA, NHLBI Grant R01-HL159205 (to L.P.).
- **Competing Interests:** The authors declare no competing interests.

## Appendix A Supplementary Information

### A.1 Landmarks sampling pipeline

The SSA pipeline described in Section 2.3 relies on a consistent parameterization of each aortic surface. Algorithm 1 details the procedure used to extract *N_s_* × *N_θ_* landmark points per scan. At each centerline sample point, a cutting plane is defined by the local tangent vector, and the intersection curve with the aortic surface is resampled at equally spaced polar angles. The starting direction vector is propagated from one cross-section to the next to maintain angular consistency along the vessel.

#### Algorithm 1

**Aorta’s landmarks sampling**

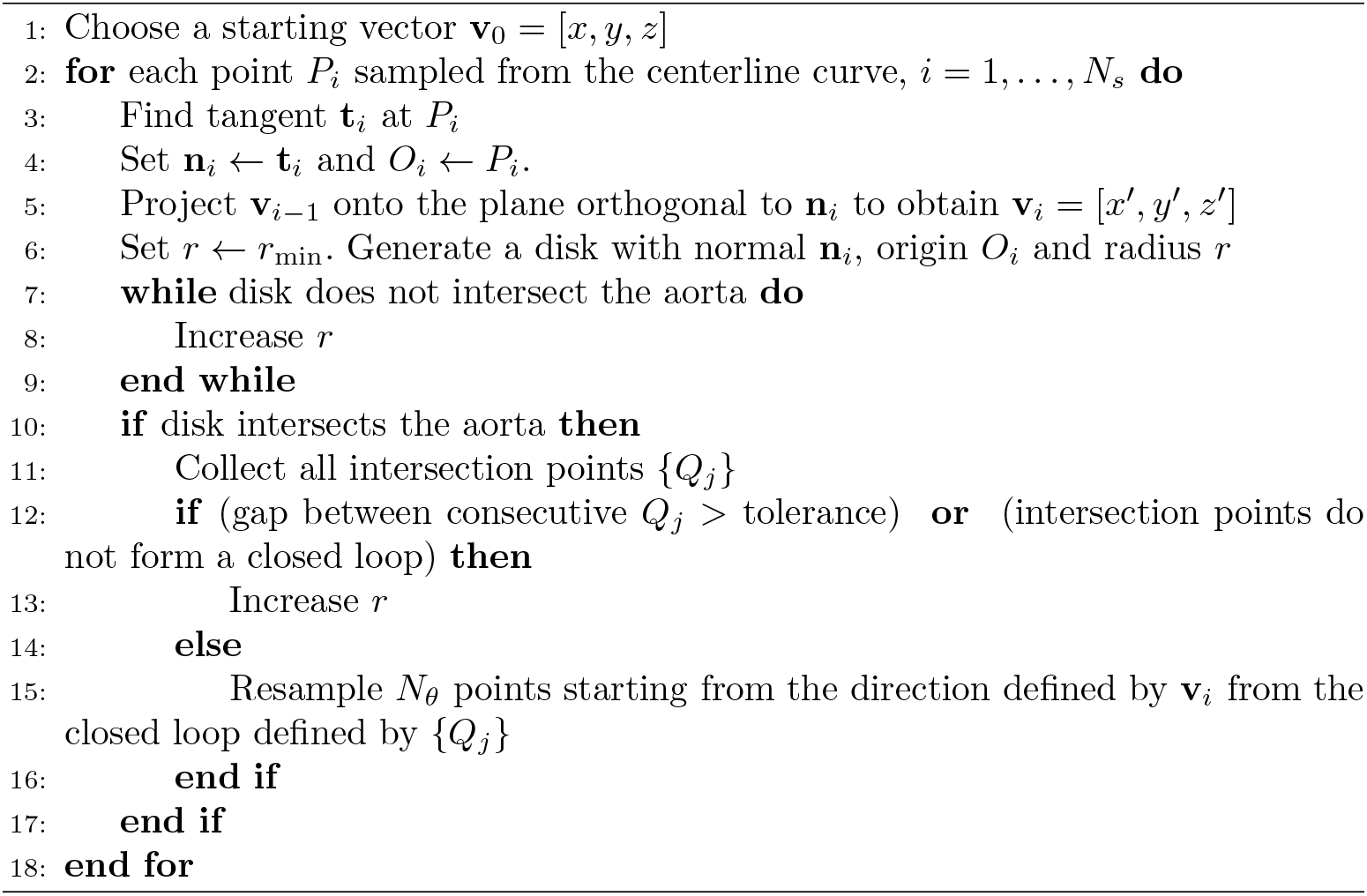

A small number of cross-sections near the aortic arch produced incomplete intersection loops due to high local centerline curvature, which can cause the tangent plane to intersect branch vessels or miss the wall entirely. While noise doesn’t contribute to the main principal components, such as PC_1_ or PC_2_, these misplaced landmark points might mix with subsequent principal components and affect classification results. For the results reported in this paper, affected landmarks are retained without manual correction; because PCA concentrates most variance in the leading components, isolated noisy landmarks have minimal impact on PC_1_ and PC_2_ but may contribute to the noise-dominated higher-order components discussed in Section 3.2.

### A.2 Supplementary Classification Results

Figure A1 shows the distribution of the weighted *F*_1_ score difference between SSA and geometric approach when applying to the same bootstrap samples drawn from the 290-scans cohort. Two GPCs are trained on either the first two principal components’ score (SSA approach) or (*R*, 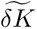) and are evaluated on out-of-bag scans. Bootstrapping allows us to compare the two approaches under repeated resampling of the cohort, thereby quantifying the variability and uncertainty in their relative performance. On average, the geometric approach performs better than SSA approach, but this difference does not reach statistical significance, as the 95% confidence interval of the performance difference includes zero.

**Fig. A1.**
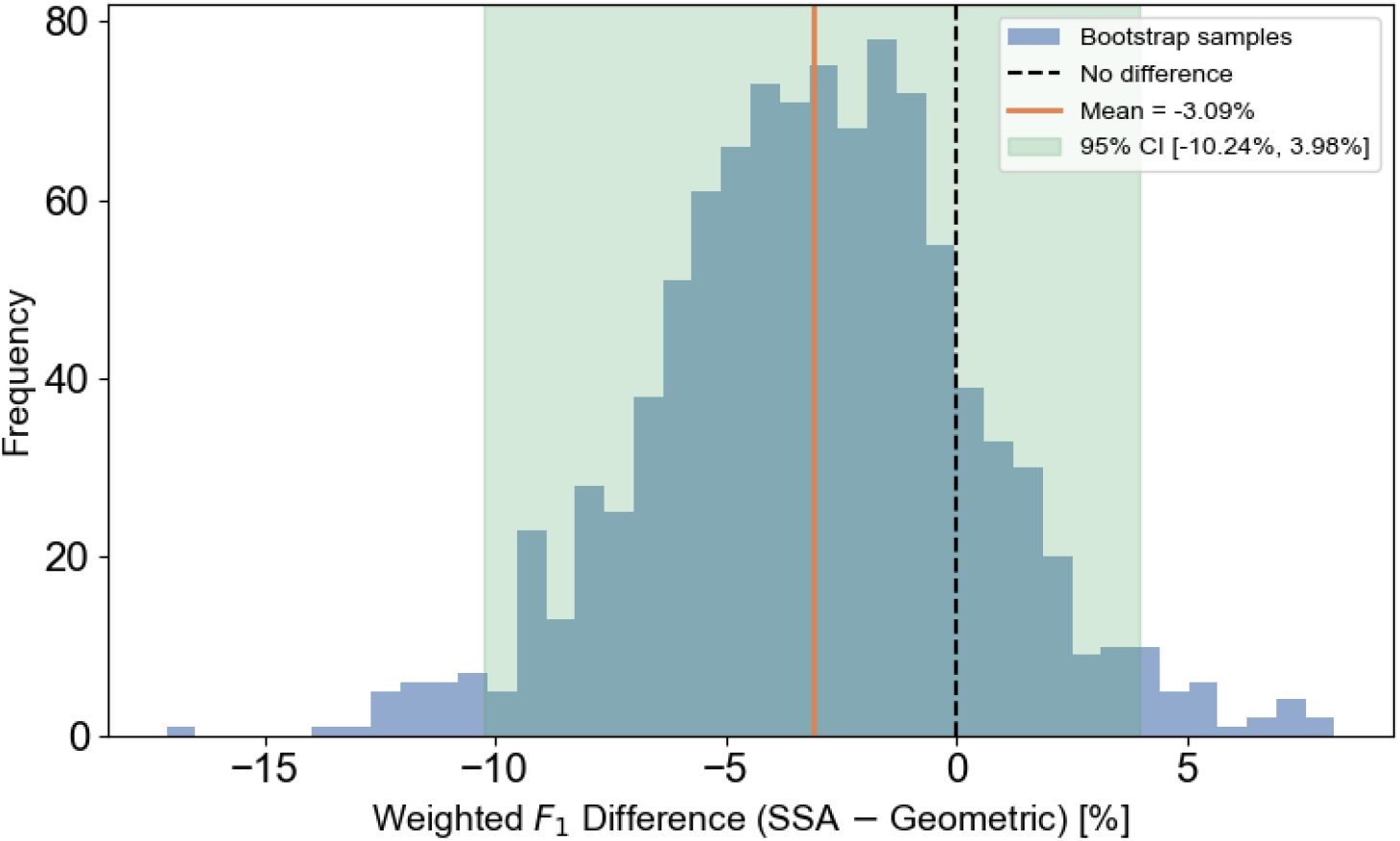
Bootstrap distribution (1000 samples) of the GPC-predicted weighted *F*_1_ score difference between SSA (2 features) and geometric approach, resampled from the 290-scan cohort. Positive values mean that SSA approach achieves a higher *F*_1_ score. The dashed black line indicates the point where both approaches achieve equal performance, the orange vertical line indicates the mean difference, and the green shaded region shows 95% confidence interval. On average, the geometric approach achieves better *F*_1_ score than SSA. However, this gain in performance by the geometric approach does not reach *p <* 0.05 significance level, as the 95% confidence interval passed the dashed black line.

Figure A2 presents the same head-to-head comparison shown in Figure 4, but using multinomial logistic regression in place of Gaussian process classification. With a linear decision boundary, the two approaches yield similar weighted *F*_1_ scores (85.3% for SSA vs. 86.5% for the geometric descriptors), confirming that the performance gap between the methods is modest when classifier flexibility is limited. The advantage of the geometric feature space becomes more pronounced under the nonlinear GPC model (Figure 4), where the physically grounded axes allow the classifier to carve out distinct outcome regions that align with anatomical intuition.

**Fig. A2.**
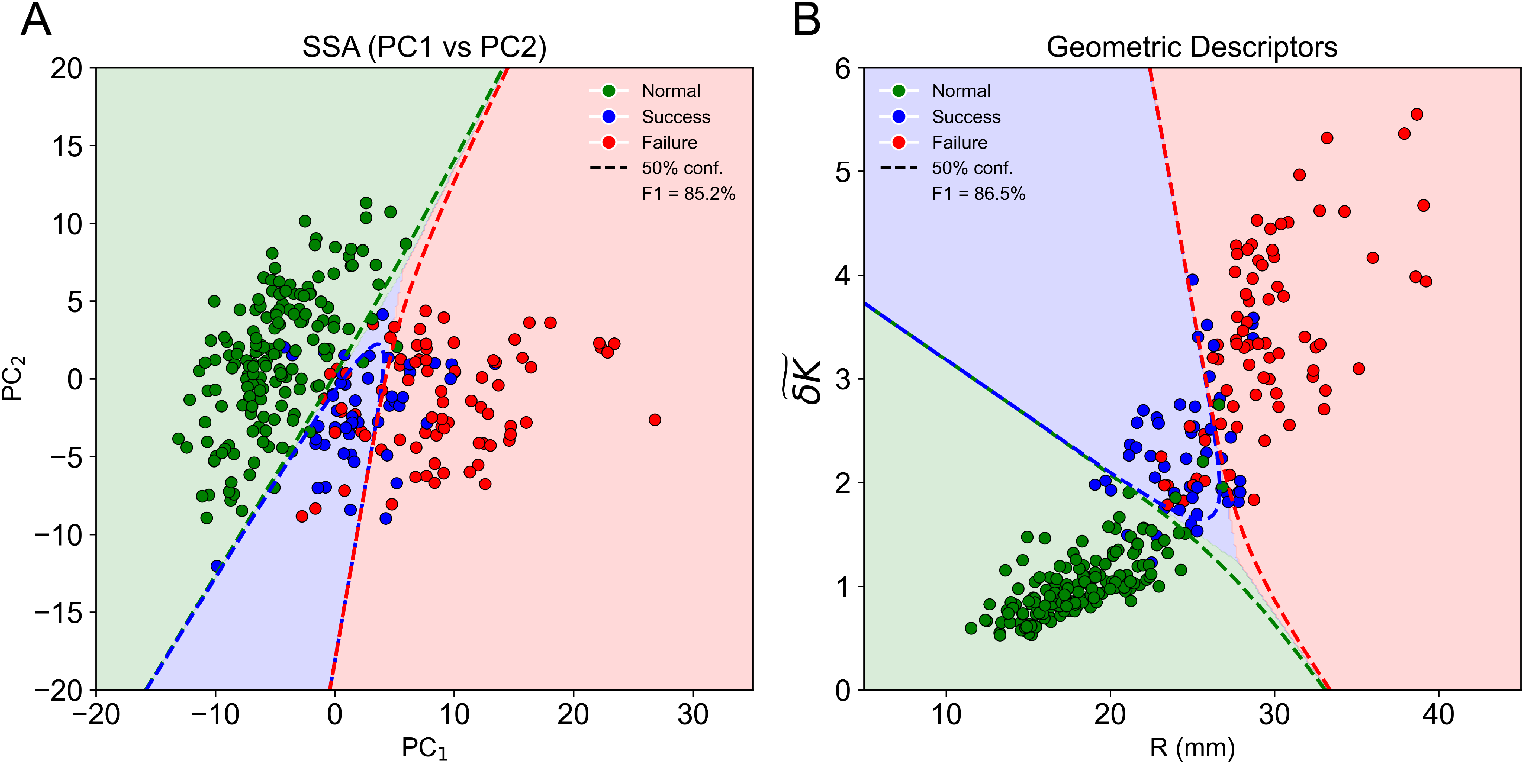
Multinomial logistic regression decision boundaries. **(A)** SSA feature space (PC_1_, PC_2_) with linear decision boundaries from multinomial LR (weighted *F*_1_ = 85.2%). Shaded regions indicate predicted class territories; dashed lines denote the 50% probability contour for each class. **(B)** Geometric descriptor feature space (*R* vs. 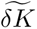) with multinomial LR boundaries (weighted *F*_1_ = 86.5%). The linear model in the geometric space yields comparable discrimination to SSA, with boundaries that are directly interpretable in terms of aortic size and surface irregularity. Green: Normal; blue: TEVAR Success; red: TEVAR Failure.

### A.3 Mesh Alignment Quality

Reliable SSA requires that inter-subject differences in the landmark representation reflect genuine anatomical variation rather than residual misalignment. Figure A3 provides a more detailed view of the registration quality summarized in Figure 2**(A)**. At the tightest tolerance (*τ* = 0.01), off-diagonal Surface DICE scores are uniformly low, as expected given true anatomical diversity across subjects. As the tolerance increases to *τ* = 0.1, nearly all pairwise scores exceed 0.9, confirming that the Procrustes-based pipeline brings meshes into close spatial agreement at the scale relevant to our landmark sampling (cross-sectional spacing on the order of centimeters).

**Fig. A3.**
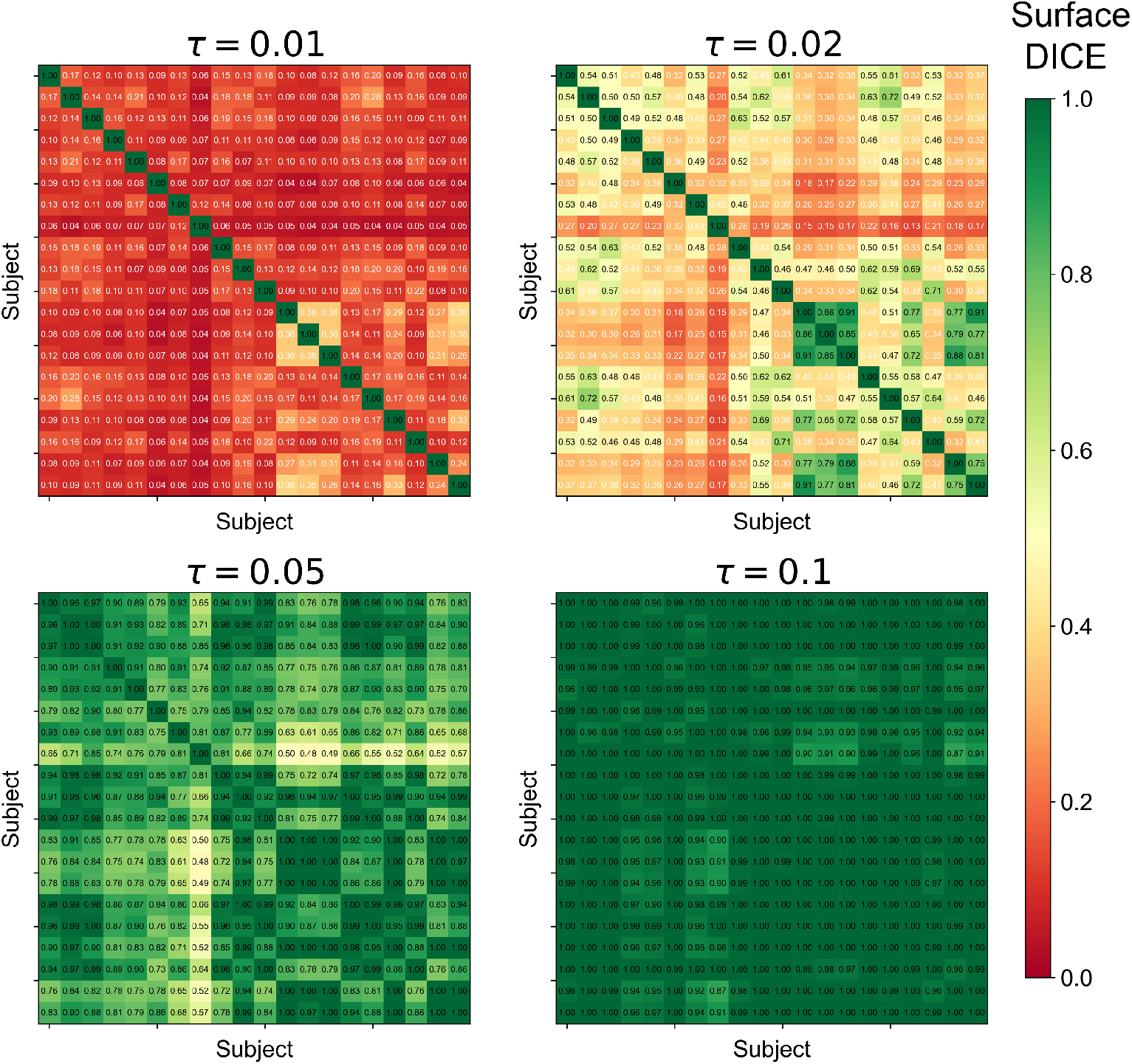
Pairwise Surface DICE heatmaps across tolerance thresholds. Each panel shows the pairwise Surface DICE coefficient for a random subset of 20 aligned subjects at tolerance *τ* ∈ {0.01, 0.02, 0.05, 0.1}. Meshes are subsampled to 1 000 points each. At tight tolerances (*τ* = 0.01), off-diagonal scores are uniformly low, reflecting genuine inter-subject anatomical differences. As *τ* increases, the heatmap transitions toward uniformly high agreement (*τ* = 0.1, nearly all entries *≥* 0.9), indicating that the Procrustes alignment successfully registers aortic surfaces to within this spatial tolerance. The diagonal is identically 1.0 by definition.

### A.4 Principal Component Space Visualization

Figure A4 displays all six pairwise projections of the first four principal components extracted by the SSA pipeline. Consistent with the classification results in Section 3.1, the PC_1_–PC_2_ plane provides the clearest separation between normal and pathological groups, while higher-order components show substantial class overlap. This visualization motivates the focus on two-dimensional feature spaces throughout the main text and supports the observation in Section 3.2 that retaining additional principal components beyond *N_p_* ≈ 3 contributes little generalizable discriminative information.

**Fig. A4.**
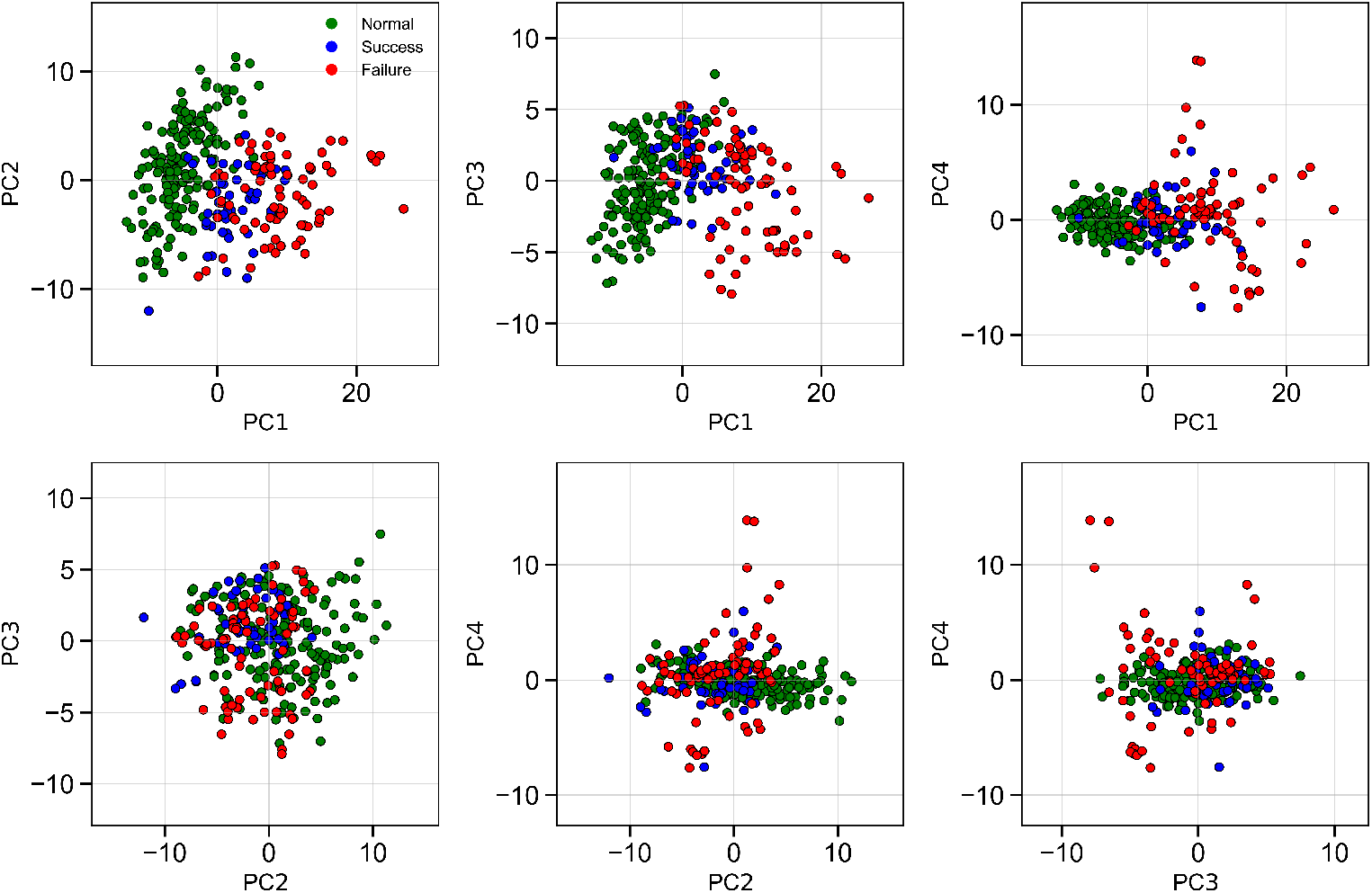
Pairwise scatter plots of the first four principal components. All six pairwise projections of PC_1_ through PC_4_ from the SSA pipeline, colored by outcome class (green: Normal; blue: TEVAR Success; red: TEVAR Failure). The PC_1_–PC_2_ plane provides the strongest visual separation between normal and pathological groups, consistent with the dominance of these components in explaining overall data variance. Higher-order components (PC_3_, PC_4_) show substantially more class overlap, indicating that additional PCs contribute limited discriminative information.

### A.5 Correlation Between Principal Components and Geometric Descriptors

A central question raised in Section 4.1 is whether the SSA feature space encodes the same anatomical information captured by the geometric descriptors. Figures A5 and A6 address this by plotting each of the first four principal components against mean aortic radius and 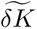, respectively.

PC_1_ is strongly correlated with both *R* (*r* = 0.80) and 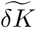 (*r* = 0.83), confirming that the dominant mode of shape variation captured by PCA is largely co-linear with the size–shape axis that defines the geometric feature space. PC_2_ shows moderate inverse correlations with both descriptors (*r* = −0.44 for *R*, *r* = −0.36 for 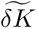), suggesting that it captures a mixture of size and shape variation that cannot be cleanly attributed to either quantity alone. PC_3_ and PC_4_ are effectively uncorrelated with both geometric descriptors, indicating that higher-order shape modes encode anatomical variation—or noise—that is orthogonal to the clinically relevant size and curvature heterogeneity captured by *R* and 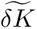.

Taken together, these results demonstrate that the discriminative information in the SSA manifold is largely redundant with the two geometric descriptors, which carry the additional advantage of direct physical interpretability.

**Fig. A5.**
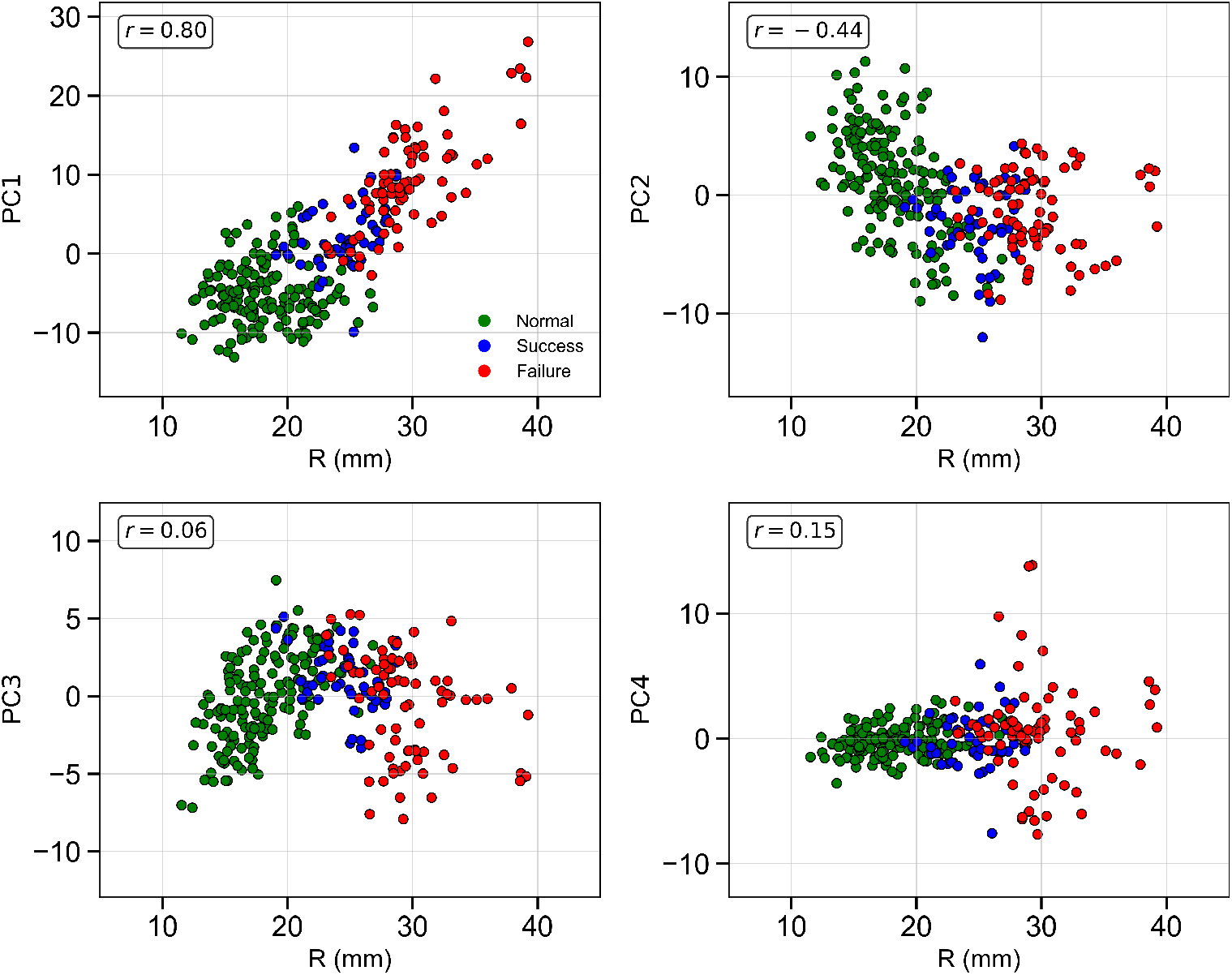
Correlation between SSA principal components and mean aortic radius. Scatter plots of mean aortic radius (mm) versus each of the first four principal components, colored by outcome class. Pearson correlation coefficients are shown inset. PC_1_ is strongly correlated with mean radius (*r* = 0.80), confirming that the leading mode of shape variation captured by SSA predominantly reflects overall aortic size. PC_2_ shows a moderate inverse correlation (*r* = *−*0.44), while PC_3_ and PC_4_ are effectively uncorrelated (*r* = 0.06 and *r* = 0.15, respectively).

**Fig. A6.**
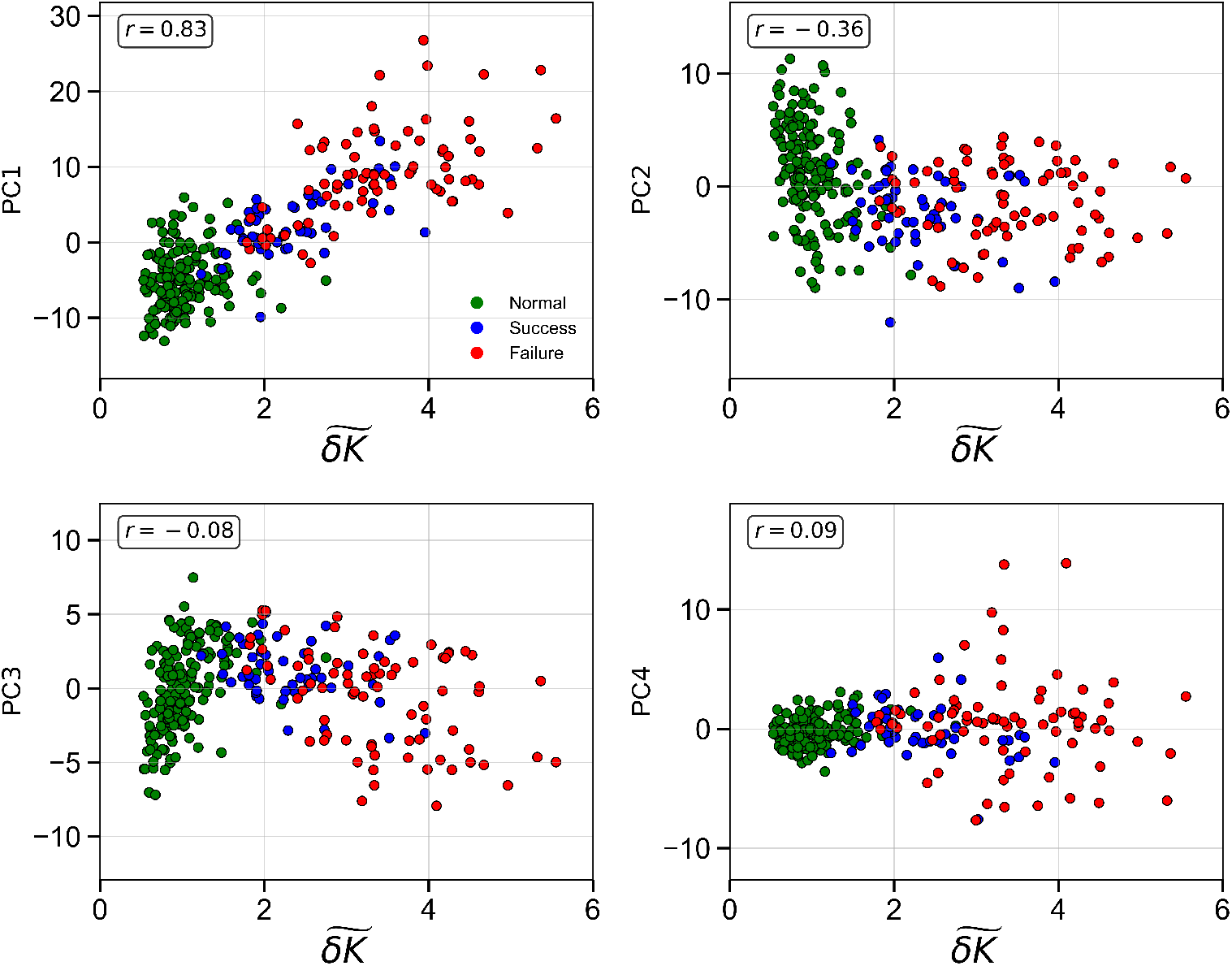
Correlation between SSA principal components and Gaussian curvature fluctuation. Scatter plots of normalized Gaussian curvature fluctuation (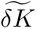) versus each of the first four principal components, colored by outcome class. PC_1_ exhibits a strong positive correlation with 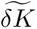 (*r* = 0.83), indicating that the dominant mode of variation tracked by SSA is roughly co-linear with the curvature-based geometric descriptor. PC_2_ shows a moderate negative correlation (*r* = *−*0.36), while PC_3_ and PC_4_ are essentially uncorrelated (*r* = *−*0.08 and *r* = 0.09). Together with Figure A5, these results demonstrate that much of the discriminative information in the SSA manifold is redundant with the two geometric descriptors (*R* and 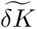), which carry the additional advantage of direct clinical interpretability.

## Notes

### Competing Interest Statement

The authors have declared no competing interest.

### Author Declarations

This study was conducted in accordance with relevant laws, institutional guidelines, and the ethical principles outlined in the Declaration of Helsinki. Patient imaging data were obtained through the Human Imaging Research Office (HIRO) at the University of Chicago under Institutional Review Board (IRB) approvals IRB20-0653 (approved 7/15/2020) and IRB21-0299 (approved 3/24/2021).

## References

[1] Isselbacher, E.M., Preventza, O., Hamilton Black III, J., Augoustides, J.G., Beck, A.W., Bolen, M.A., Braverman, A.C., Bray, B.E., Brown-Zimmerman, M.M., Chen, E.P., Collins, T.J., DeAnda, A. Jr, Fanola, C.L., Girardi, L.N., Hicks, C.W., Hui, D.S., Schuyler Jones, W., Kalahasti, V., Kim, K.M., Milewicz, D.M., Oderich, G.S., Ogbechie, L., Promes, S.B., Gyang Ross, E., Schermerhorn, M.L., Singleton Times, S., Tseng, E.E., Wang, G.J., Woo, Y.J., Peer Review Committee Members: 2022 ACC/AHA Guideline for the Diagnosis and Management of Aortic Disease: A Report of the American Heart Association/American College of Cardiology Joint Committee on Clinical Practice Guidelines. Circulation 146(24), 334–482 (2022) 10.1161/CIR.0000000000001106

[2] Geronzi, L., Martínez, A., Rochette, M., Yan, K., Bel-Brunon, A., Haigron, P., Escrig, P., Tomasi, J., Daniel, M., Lalande, A., Lin, S., Marín-Castrillón, D., Bouchot, O., Porterie, J., Valentini, P., Biancolini, M.: Computer-aided shape features extraction and regression models for predicting the ascending aortic aneurysm growth rate. Computers in Biology and Medicine 162, 107052 (2023) 10.1016/j.compbiomed.2023.107052

[3] Bruse, J.L., Khushnood, A., McLeod, K., Biglino, G., Sermesant, M., Pennec, X., Taylor, A., Hsia, T., Schievano, S.: How successful is successful? Aortic arch shape after successful aortic coarctation repair correlates with left ventricular function. Journal of Thoracic and Cardiovascular Surgery 153(2), 418–427 (2017) 10.1016/j.jtcvs.2016.09.018

[4] Lee, K., Zhu, J., Shum, J., Zhang, Y., Muluk, S.C., Chandra, A., Eskandari, M.K., Finol, E.A.: Surface Curvature as a Classifier of Abdominal Aortic Aneurysms: A Comparative Analysis. Ann. Biomed. Eng. 41(3), 562–576 (2013) 10.1007/s10439-012-0691-4

[5] Carmo, M.P.: Differential Geometry of Curves and Surfaces. Prentice–Hall, Englewood Cliffs, NJ (1976)

[6] Khabaz, K., Yuan, K., Pugar, J., Jiang, D., Sankary, S., Dhara, S., Kim, J., Kang, J., Nguyen, N., Cao, K., Washburn, N., Bohr, N., Lee, C.J., Kindlmann, G., Milner, R., Pocivavsek, L.: The geometric evolution of aortic dissections: Predicting surgical success using fluctuations in integrated Gaussian curvature. PLoS Comput. Biol. 20(2), 1011815 (2024) 10.1371/journal.pcbi.1011815

[7] Pugar, J.A., Jiang, D., Kim, J., Pocivavsek, L.: Shape and Scale in Quantifying Aortic Morphology Evolution and Chronicity. Cardiovasc. Eng. Technol. (2026) 10.1007/s13239-026-00827-z

[8] Cosentino, F., Raffa, G., Gentile, G., Agnese, V., Bellavia, D., Pilato, M., Pasta, S.: Statistical Shape Analysis of Ascending Thoracic Aortic Aneurysm: Correlation between Shape and Biomechanical Descriptors. Journal of Personalized Medicine 10(2), 28 (2020) 10.3390/jpm10020028

[9] Shehata, N., Elsawy, A., Nagy, M., ElMahdy, M., Ali, M., Romeih, S., Aguib, H., Yacoub, M., Glocker, B.: A Comprehensive Pipeline for Aortic Segmentation and Shape Analysis. arXiv (2025) 10.48550/arXiv.2509.09718

[10] Veldhuizen, W.A., Schuurmann, R., IJpma, F., Kropman, R., Antoniou, G., Wolterink, J., Vries, J.D.: A Statistical Shape Model of the Morphological Variation of the Infrarenal Abdominal Aortic Aneurysm Neck. Journal of Clinical Medicine 11(6), 1687 (2022) 10.3390/jcm11061687

[11] Aljassam, Y., Sophocleous, F., Bruse, J.L., Schot, V., Caputo, M., Biglino, G.: Machine Learning and Statistical Shape Modelling Methodologies to Assess Vascular Morphology before and after Aortic Valve Replacement. Journal of Clinical Medicine 13(15), 4577 (2024) 10.3390/jcm13154577

[12] Hermida, U., Poppel, M.V., Lloyd, D., Steinweg, J., Vigneswaran, T., Simpson, J., Razavi, R., Vecchi, A., Pushparajah, K., Lamata, P.: Learning the Hidden Signature of Fetal Arch Anatomy: a Three-dimensional Shape Analysis in Suspected Coarctation of the Aorta. Journal of Cardiovascular Translational Research 16, 738–747 (2022) 10.1007/s12265-022-10335-9

[13] Wang, Y., Yang, Q., Li, J., Wang, K., Tang, M.: Automatic measurement of proximal femoral morphological parameters using point cloud semantic segmentation technology. Sci. Rep. 15(1), 12551 (2025) 10.1038/s41598-025-94310-9

[14] Zeng, Y., Sun, Z., Wang, M., Li, Z., Liu, A., Pan, M., Zhao, H., Li, Y.: Expanding point cloud statistical shape model applications: Generalized vascular modeling for population-level hemodynamic simulations. Comput. Methods Programs Biomed. 269(108924), 108924 (2025) 10.1016/j.cmpb.2025.108924

[15] Bruse, J.L., McLeod, K., Biglino, G., Ntsinjana, H.N., Capelli, C., Hsia, T.-Y., Sermesant, M., Pennec, X., Taylor, A.M., Schievano, S., Modeling of Congenital Hearts Alliance (MOCHA) Collaborative Group: A statistical shape modelling framework to extract 3D shape biomarkers from medical imaging data: assessing arch morphology of repaired coarctation of the aorta. BMC Med. Imaging 16(1), 40 (2016) 10.1186/s12880-016-0142-z

[16] Sophocleous, F., Biffi, B., Milano, E.G., Bruse, J., Caputo, M., Rajakaruna, C., Schievano, S., Emanueli, C., Bucciarelli-Ducci, C., Biglino, G.: Aortic morphological variability in patients with bicuspid aortic valve and aortic coarctation. Eur. J. Cardiothorac. Surg. 55(4), 704–713 (2019) 10.1093/ejcts/ezy339

[17] Pearson, K.: LIII. on lines and planes of closest fit to systems of points in space. Lond. Edinb. Dublin Philos. Mag. J. Sci. 2(11), 559–572 (1901) 10.1080/14786440109462720

[18] Zhang, M., Golland, P.: Statistical shape analysis: From landmarks to diffeomorphisms. Medical Image Analysis 33, 155–158 (2016) 10.1016/j.media.2016.06.025. 20th anniversary of the Medical Image Analysis journal (MedIA)

[19] Gao, Y., Riklin-Raviv, T., Bouix, S.: Shape Analysis, A Field in Need of Careful Validation. Hum. Brain Mapp. 35(10), 4965–4978 (2014) 10.1002/hbm.22525

[20] Sukno, F.M., Frangi, A.F.: Reliability Estimation for Statistical Shape Models. IEEE Transactions on Image Processing 17(12), 2442–2455 (2008) 10.1109/TIP.2008.2006604

[21] NUREA: PraevAorta: Advanced Aortic Image Analysis Software. https://www.nurea.com/praevaorta (2025)

[22] Rusinkiewicz, S.: Estimating curvatures and their derivatives on triangle meshes. In: Proceedings. 2nd International Symposium on 3D Data Processing, Visualization and Transmission, 2004. 3DPVT 2004, pp. 486–493. IEEE,(2004). 10.1109/TDPVT.2004.1335277

[23] Wiputra, H., Matsumoto, S., Wagenseil, J.E., Braverman, A.C., Voeller, R.K., Barocas, V.H.: Statistical shape representation of the thoracic aorta: accounting for major branches of the aortic arch. Computer Methods in Biomechanics and Biomedical Engineering 26(13), 1557–1571 (2022) 10.1080/10255842.2022.2128672

[24] Pedregosa, F., Varoquaux, G., Gramfort, A., Michel, V., Thirion, B., Grisel, O., Blondel, M., Prettenhofer, P., Weiss, R., Dubourg, V., Vanderplas, J., Passos, A., Cournapeau, D., Brucher, M., Perrot, M., Duchesnay, E.: Scikit-learn: Machine Learning in Python. Journal of Machine Learning Research 12, 2825–2830 (2011)

[25] Nikolov, S., Blackwell, S., Zverovitch, A., Mendes, R., Livne, M., De Fauw, J., Patel, Y., Meyer, C., Askham, H., Romera-Paredes, B., Kelly, C., Karthikesalingam, A., Chu, C., Carnell, D., Boon, C., D’Souza, D., Moinuddin, S.A., Garie, B., McQuinlan, Y., Ireland, S., Hampton, K., Fuller, K., Montgomery, H., Rees, G., Suleyman, M., Back, T., Hughes, C.O., Ledsam, J.R., Ronneberger, O.: Clinically Applicable Segmentation of Head and Neck Anatomy for Radiotherapy: Deep Learning Algorithm Development and Validation Study. J. Med. Internet Res. 23(7), 26151 (2021) 10.2196/26151

[26] Desai, N., Tohill, B., Matsumura, J., Cambria, R.P.: A Multicenter clinical Trial on the Five-Year Outcomes Following Treatment of Acute, Complicated Type B Aortic Dissection with a Conformable Stent Graft. Ann. Vasc. Surg. 113, 1–12 (2025) 10.1016/j.avsg.2024.12.008

